# Predicting the Epidemic Curve of the Coronavirus (SARS-CoV-2) Disease (COVID-19) Using Artificial Intelligence

**DOI:** 10.1101/2020.04.17.20069666

**Authors:** László Róbert Kolozsvári, Tamás Bérczes, András Hajdu, Rudolf Gesztelyi, Attila Tiba, Imre Varga, Ala’a B. Al-Tammemi, Gergő József Szőllősi, Szilvia Harsányi, Szabolcs Garbóczy, Judit Zsuga

## Abstract

The aim of our study was to predict the epidemic curves (daily new cases) of COVID-19 pandemic using Artificial Intelligence (AI)-based Recurrent Neural Networks (RNNs), then to compare and validate the predicted models with the observed data. We used the publicly available datasets from the World Health Organization and Johns Hopkins University to create a training dataset, then we used RNNs with gated recurring units (Long Short-Term Memory) to create two prediction models. Information collected in the first t time-steps were aggregated with a fully connected (dense) neural network layer and a consequent regression output layer to determine the next predicted value. We also used Root Mean Squared Logarithmic Errors (RMSLE) to compare the predicted models with the observed data. The result of our study underscores that the COVID-19 pandemic is a propagated source epidemic, therefore repeated peaks on the epidemic curve are to be anticipated. Besides, the errors between the predicted and validated data and trends seems to be low. The influence of this pandemic is significant worldwide and has already impacted our daily life. Decision makers must be aware, that even if strict public health measures are executed and sustained, future peaks of infections are possible.

## 1. INTRODUCTION

### 1.1 Coronavirus

High and low pathogenic species may be distinguished within the coronavirus family, with the former including 4 viruses that are responsible for 10-30% of mild upper respiratory diseases (e.g. common cold), and the latter known to cause a more severe form of acute lung injury: SARS-CoV (Severe Acute Respiratory Syndrome) and MERS-CoV (Middle East Respiratory Syndrome) [1]. SARS-CoV originated in Guangdong Province, China, and started to spread in 2002, causing over 8,000 cases in 29 countries around the world, with a crude fatality rate of 10% [2–4]. The virus has spread to Hong Kong in 2003 causing an outbreak of severe acute respiratory syndrome (SARS). A novel coronavirus was isolated and was suggested to be the primary cause of the infections [5]. A few years later, in 2007 Cheng et. al issued a warning that “the presence of a large reservoir of SARS-CoV-like viruses in horseshoe bats, together with the culture of eating exotic mammals in southern China, is a time bomb” [4]. MERS-CoV began spreading in Saudi Arabia in 2012 and, to the time of writing this, has led to a total of 2519 laboratory-confirmed cases in several countries around the world [6,7]. Its case-fatality rate reached 37.1% over the course of the past 8 years [7].

### 1.2 Coronavirus Disease – 2019 (COVID-19)

The current form of the Severe Acute Respiratory Syndrome noted as COVID-19, is caused by a new variant of formerly known highly pathogenic *Coronaviridae*. The infection allegedly began to spread from a market in Wuhan, the capital of Hubei province, China, at the end of 2019 [8,9]. Early genome sequencing has found that the new virus, which was named SARS-CoV-2 by the International Committee on Taxonomy of Viruses, showed a 79.6% homology with SARS-CoV, and has 96% sequence identity with bat coronavirus suggesting a common origin from SARSr-CoV (severe acute respiratory syndrome-related coronavirus). Also, the suspected host was thought to be a bat species, *Rhinolophus affinis* (a horseshoe bat), but the SARS-COV-2 probably needs an intermediate host [9,10].

Symptoms associated with the COVID-19 include fever (83%), cough (82%), shortness of breath (31%), muscle aches (11%), confusion (9%), headache (8%), sore throat (5%), runny nose, chest pain, diarrhea, nausea and vomiting [11]. According to a meta-analysis that compiled data from more than 50 000 patients, the incidence of fever (0.891, 95% confidence interval (CI): [0.818; 0.945]) and cough (incidence of 0.722, 95% CI: [0.657; 0.782]) were the highest, respectively, followed by muscle soreness and fatigue [12]. The incubation period of the virus was estimated to be between 1-14 days (5 days on average) [13]. There is no definite data concerning the transmissibility of the virus. However, several transmission routes have been identified including respiratory droplets/aerosols, direct contact with virally-contaminated objects, and possibly fecal-oral transmission [14].

It seems probable that those with a fulminant disease are most infectious, but reports have identified asymptomatic and pre-symptomatic virus shedding as well. There is also a lack of definite data regarding tertiary and quaternary spreading among humans, but it seems probable that the person who has been exposed to the infection has acquired some (at least temporary) immunity [15]. According to data from the World Health Organization (WHO), there were 91,816,091 confirmed cases and 1,986,871 fatalities globally as of January 15, 2021, which corresponds to a case-fatality rate of 2.164% [16].

R0, the basic reproduction number, denoting the transmissibility of a virus that indicates the average number of new infections induced by an infectious person in a susceptible, infection naïve population. The transmissibility of the virus was apparently underestimated initially by the WHO with R0 suggested to range between 1.4 and 2.5. More recent reports have indicated higher R0 values around 3 (with the mean and median R0 for published estimates being 3.28 and 2.79, respectively) [13,17].

### 1.3 The Daily Number of Newly Diagnosed Infections - Epidemic Curves

The initial epidemic curves of the COVID-19 outbreak from Hubei, China showed a mixed pattern, indicating that early cases were likely from a continuous common source e.g., from several zoonotic events in Wuhan, followed by secondary and tertiary transmission providing a propagated source for the later cases [18].

The propagated (or progressive source) epidemic curve visualizes the spread of an infectious agent that may be transmitted from human to human starting from with a single index case, that continues to further infect other individuals. This shows up as a series of peaks on the epidemic curve, that starts with the index case, followed by successive waves of the infection set apart with respect to the incubation period of the pathogen. The waves continue to follow each other, until appropriate mitigation measures, prevention, or treatment are implemented, or the pool of the susceptible population becomes infected. This is a theoretic curve, that is generally influenced by lots of other factors [18].

Several studies investigated the impact of different interventions with respect to minimizing contact rates in the population in order to slow the infection spread, minimize COVID-19 mortality rates and health care utilization, or to suppress the epidemic per se. Flattening the curve by reducing peak incidence may limit overall case fatality rates. Nevertheless, most of the forecasts and simulations thus far started out from Bell-shaped curves, that fail to account for the progressive nature of the current outbreak given the known secondary, tertiary even quaternary transmissibility of the virus. Taking this into account it is suggested that the number of cases will rise once again after pandemic control measures are no longer in effect [19].

### 1.4 Prediction

There are different mathematical models that may demonstrate and predict the dynamics of different infectious diseases [20]. These models, used to simulate the dynamics of infectious diseases, may be based on statistical, mathematical, empirical, or machine learning methods [21].

The first attempts to use Artificial Intelligence (AI) in medicine were made in the 1970s. Initially, AI was used to implement programs to help clinical decision-making, but to date, its use is gaining more and more widespread acceptance in biomedical sciences [22]. One class of AI, a form of artificial neural networks, the *Recurrent Neural Networks (RNNs) with Long Short-Term Memory (LSTM)* were previously used to model and forecast the influenza epidemic, with strong competitiveness and reliable results [23–25].

### 1.5 Aim of the Study

The current study aimed to use the publicly available official COVID-19 data as a training dataset, to predict the possible outcomes of the COVID-19 pandemic (epidemic curve of new cases) using AI-based RNNs, and further, to compare the predictions with the observed data.

## 2. MATERIALS AND METHODS

### 2.1 Data

We used the publicly available datasets from the WHO and Johns Hopkins University for the following countries to create the ***training dataset***: Austria, Belgium, China (Hubei), Czech Republic, France, Germany, Hungary, Iran, Italy, Netherlands, Norway, Portugal, Slovenia, Spain, Switzerland, United Kingdom (UK) and the United States of America (USA) [15,26]. Given that most infected people in China were from Hubei province, only data from that province was included. For each country, the date of the first reported infection was set as ***Day 1*** for the disease time scale. (**Figure 1**).

**Figure 1.**
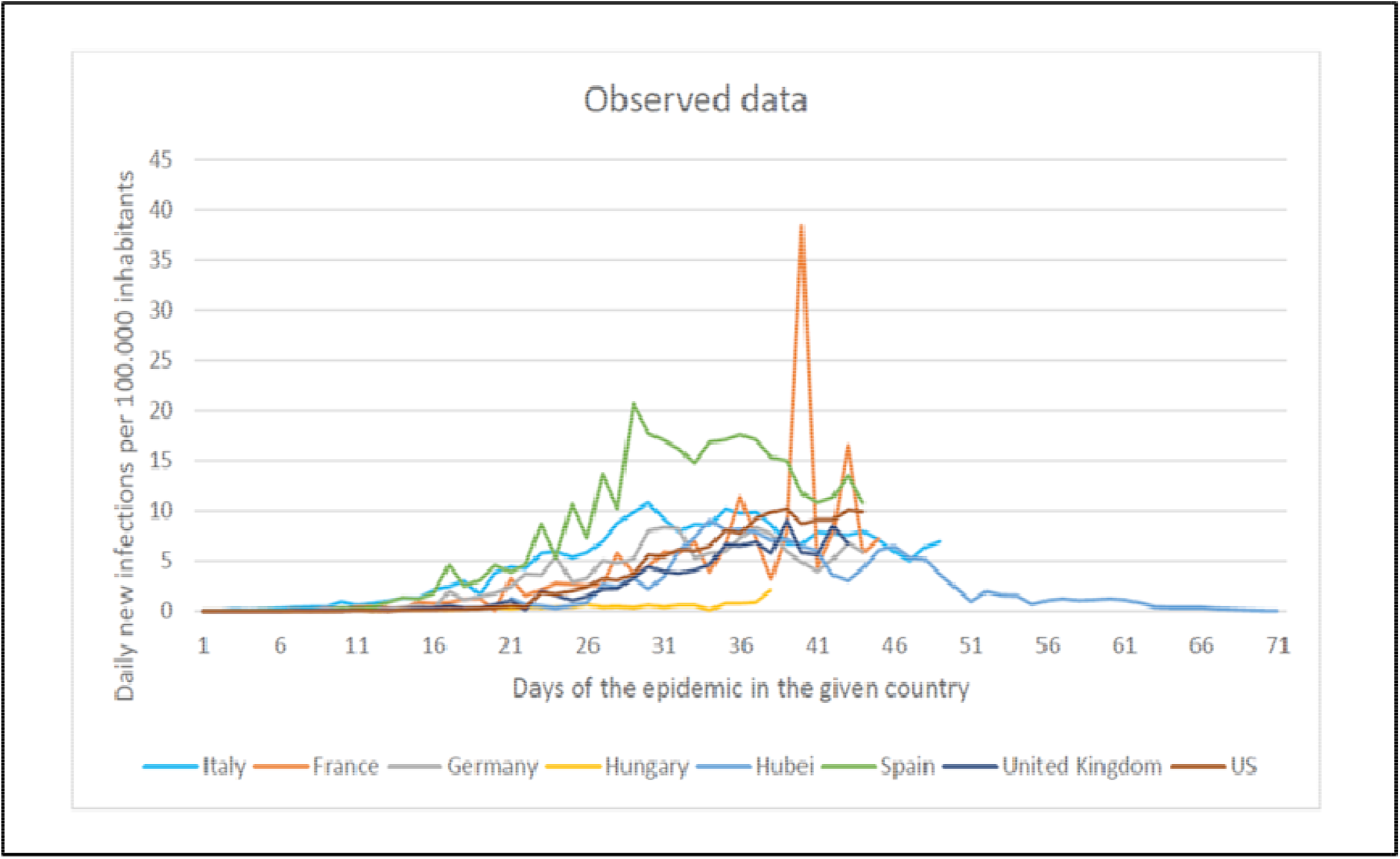
The Historical Datasets of Different Countries

When determining the date of the first illness, point-source outbreaks were omitted (e.g., those cases where single verified cases were isolated, and no further transmission has occurred). This was important to avoid distortion of the propagated epidemic curves. In Belgium, for example, the first illness occurred on February 02, 2020 and there were no further cases reported for up to 26 days. The next illness occurred on March 01, 2020. Inclusion of the early case from February would contribute to a false learning rule for the AI, hence corrupting the results. As for Hubei Province, the first officially available data is of January 22, 2020. This cannot be considered as the first day of the illness, thus the first infection was arbitrarily defined to occur on January 01, 2020. To account for the extreme variability of daily incident cases reported which probably reflects delays in reporting procedures, a moving average was used (covering 3 days) for the Hubei dataset.

Accordingly, an epidemic curve was obtained for each country with a time series where the first day denotes the day of the first confirmed case, and each successive day indicating the number of newly confirmed cases that day. To account for the country-specific differences in the size of the population, the number of daily new cases was normalized for 100.000 inhabitants in each country. The observation period varies for each country, given the difference of time elapsed since the disease initiation in that country. Accordingly, the longest time series covers the observation period of 90 days. e.g., in Hubei, with the first 22 days lacking valid data and the next 68 days having data. The shortest observation period was in Slovenia with only 30 days.

The training data set was obtained by averaging the daily incidence rates per 100 000 inhabitants across the 17 countries included, for each day in the time series. When calculating the average, missing data was left blank, i.e., NULL, e.g., countries that did not contain data for a specific day were excluded from the calculation of average. The resulting ***training data*** set is shown in **Figure 1**. It should be noted that the first part of the data set (up to the initial 30 days since **Day 1** of the epidemic) contains data for almost all the countries listed, whereas the end of the data set contains only data from Hubei. (**Figure 2**)

**Figure 2.**
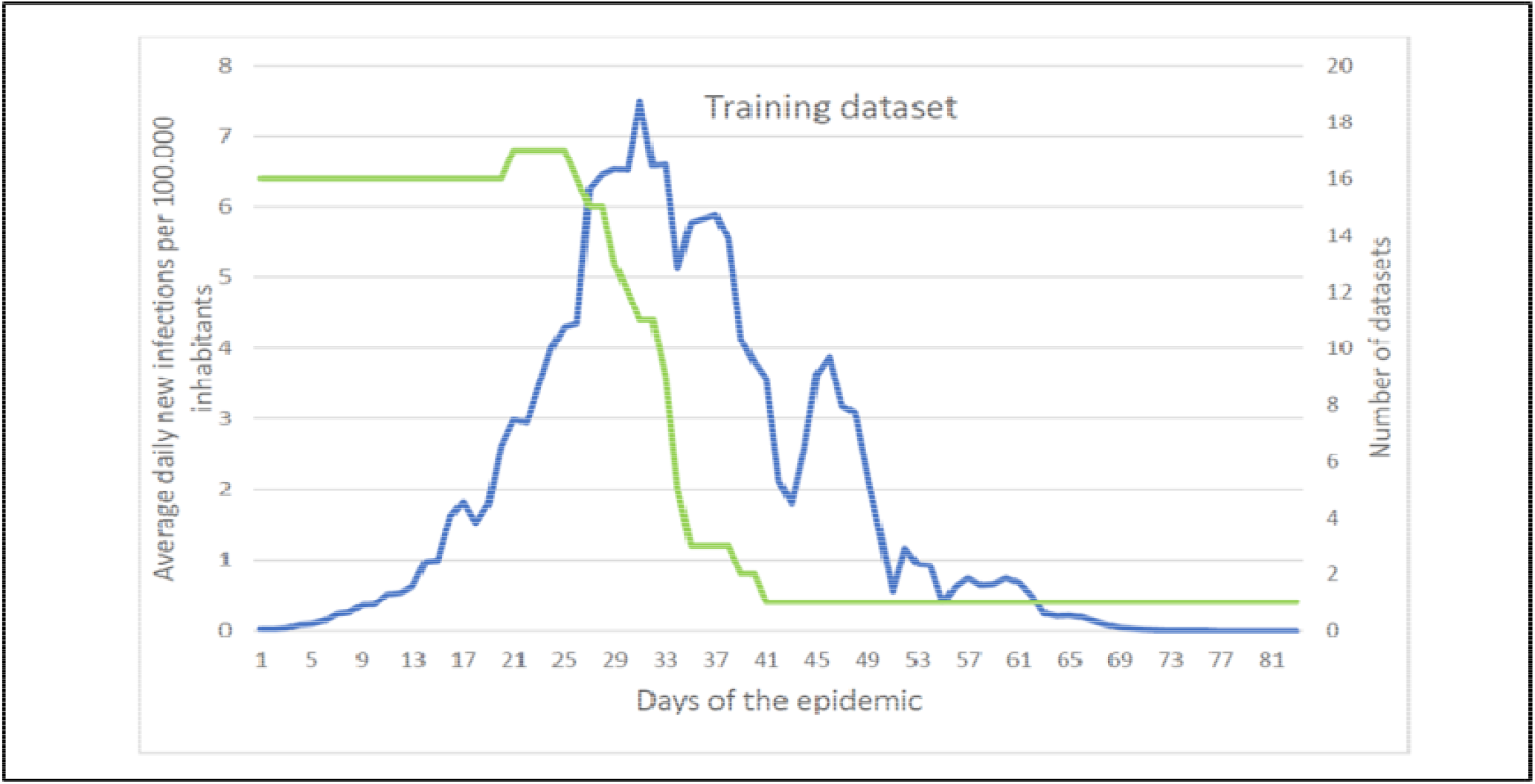
The training dataset. Average daily new infections per 100.000 inhabitants (line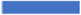) and the Number of datasets (line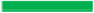)

### 2.2 RNNs-Based Model for Prediction

The state-of-the-art for time series analysis is AI-based analytic tools, which have the best prediction performance. Recurrent Neural Networks (RNNs) are specifically designed to cope with sequential input, characteristic of textual or temporal data [24]. This architecture is a neural network-based architecture, that contains hidden layers chained according to the time step, with a possibility to predict the next sequence element(s). A time series has a special temporal form, where the input to the i-th hidden layer is at the i-th time-step that has a corresponding x(i) observation. In its original form, a simple RNN tries to predict the next sequence element, however, for the purposes of the current analysis, an encoder-decoder variant is a more natural choice, similarly to machine translation [27].

For our specific scenario this means that during the encoder phase including time steps 1,…,t the RNN is fed with the already known time series data (the average of the number of new cases normalized to 100 000 inhabitants for day 1…t, respectively), followed by prediction in the decoder phase for the future time steps t+1,…,T. In our analysis, T=t+1=90 days is the longest known (Hubei) time interval. Since this covers quite a long data sequence, we have used gated recurring units (namely Long Short-Term Memory – LSTM units) in compliance with the general recommendations [25]. **Figure 3** depicts our RNN architecture showing how unknown time series elements are predicted. **Figure 3** also shows how the information collected in the first t time-steps are aggregated with a fully connected (dense) neural network layer and a consequent regression output layer to determine a predicted number of new patients as x(t+1).

**Figure 3.**
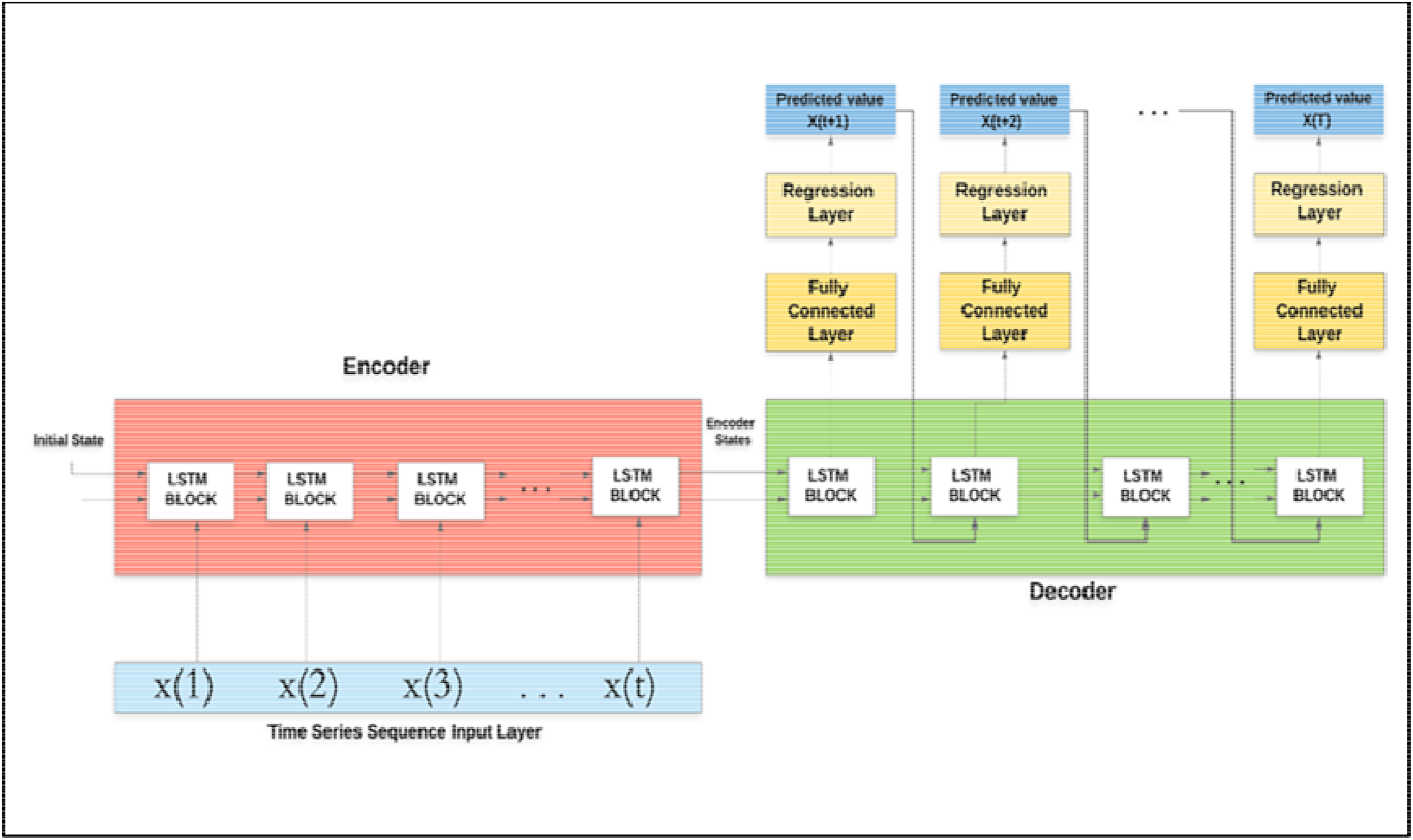
The Recurrent Neural Network (RNN) Architecture used for Prediction.

The training data was described in the previous section. To assess possible specificities regarding the countries two approaches were used for prediction:

- **Prediction 1**: An algorithm to update the training step and subsequent prediction was formulated. This update step is based on the general recommendations of transfer learning that considers the already known time interval for the given country and re-training is done in small increments of the RNN network accordingly [28]. Thus, we start predicting the first unknown element x(t+1) from the last 5% of the known data, and the same principle is applied to each subsequent element. Moreover, after each prediction step our RNN architecture was re-trained and the subsequent elements were predicted with this updated RNN.
- **Prediction 2**: We start predicting the first unknown element x(t+1) from the last known x(t), and all the subsequent elements are predicted only from the preceding ones. Here the rules depicted from the training data set are used, not retraining occurs.

The intuitive interpretations of the difference between Prediction 1 and Prediction 2 are as follows. Prediction 2 makes its predictions utilizing the information derived from the training data set, reflective of the trends in the average time series. It follows that predictions will comply primarily with the Hubei time series, especially in the far future. Therefore Prediction 2 shows the highest fidelity to the country-specific future scenario if the approach to mitigate the epidemic is similar to that in Hubei. Accordingly, this scenario is also reflective of a country-specific future state given the practices of Hubei were followed in said country. On the other hand, Prediction 1 is yielded after the neural network is retrained after any prediction, providing more valid insight into what is expected if the country goes on with the mitigation practices seen during the observation period. The architecture was trained in 250 epochs with a total number of 100 hidden LSTM layers, to prepare a bit for prediction also after T=90 days. Naturally, the length of the RNN can be freely increased later.

### 2.3 Validation

For the learning dataset, we used the data from the first pandemic wave. That is, we took the available factual data from the first case reported in a country until April 10, 2020. Based on that, we have made the above mentioned two predictions (1 and 2). Moreover, for the validation process, we used the factual data of the first wave. By country, we considered 85-90 days from the first case reported. Thus, the number of days predicted varied from country to country in the same way as for the learning dataset. The amount of Root Mean Squared Logarithmic Errors (RMSLE) was used for validation. In our analysis, the possible bias regarding the different ratios between the observed and predicted values are interpreted using the RMSLE. Let **n** be the number of days used for validation. Let **p_1i** and **p_2i** be the number of new cases per day obtained using the two prediction methods in the examined time interval and let **a_i** be the actual data for the given days. **Err1** and **Err2** will be RMSLE for Prediction 1 and Prediction 2, respectively, where:

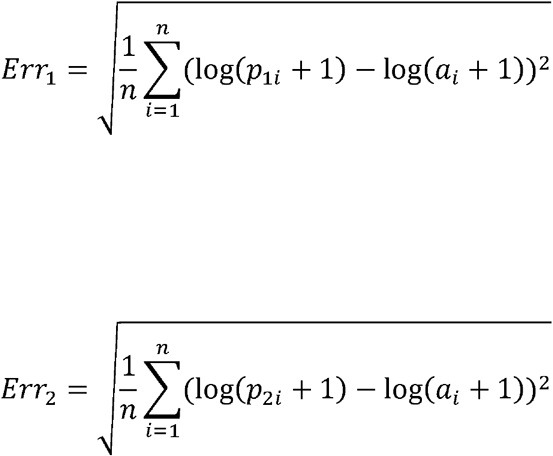

## 3. RESULTS

This section shows the outcomes for **Prediction 1** and **Prediction 2** for the individual country-level data for France, Germany, Hungary, Italy, Spain, USA, and UK (**Figures 4-10**). For each main graph, the small graph in the upper right corner contains the daily error values calculated for the predictions. The more accurate the prediction, the smaller the RMSLE error. It should be noted that if the error function is parallel to the x-axis, it means that the trend of the prediction is the same as the real trend, only at a lower or higher scale. Also, the total errors for the entire investigated period, the summarized mean of the predictions (RMSLE) by country, is shown in **Table 1**.

**Table 1.** Total RMSLE error for the entire investigated period

| **Country** | **Mean of**<br>**RMSLE of**<br>**Prediction 1** | **Mean of**<br>**RMSLE of**<br>**Prediction 2** |
| --- | --- | --- |
| Hungary | 0.06 | 0.107 |
| UK | 0.234 | 0.455 |
| Italy | 0.114 | 0.155 |
| Spain | 0.266 | 0.181 |
| Germany | 0.147 | 0.108 |
| France | 0.513 | 0.307 |
| USA | 0.216 | 0.528 |

**Figure 4.**
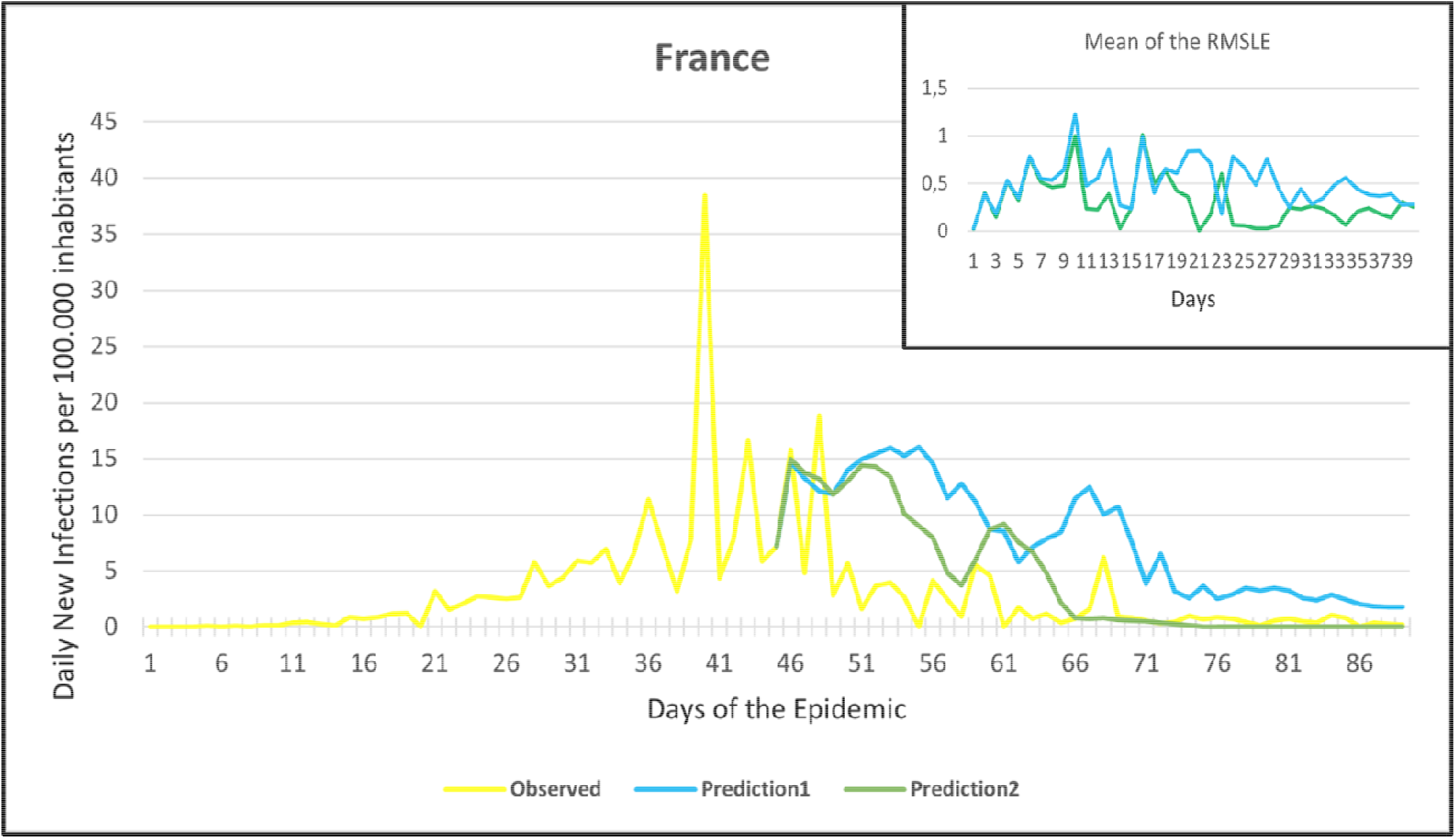
Observation and predictions for France. The small graph in the upper right corner shows the daily error values calculated for the predictions.

**Figure 5.**
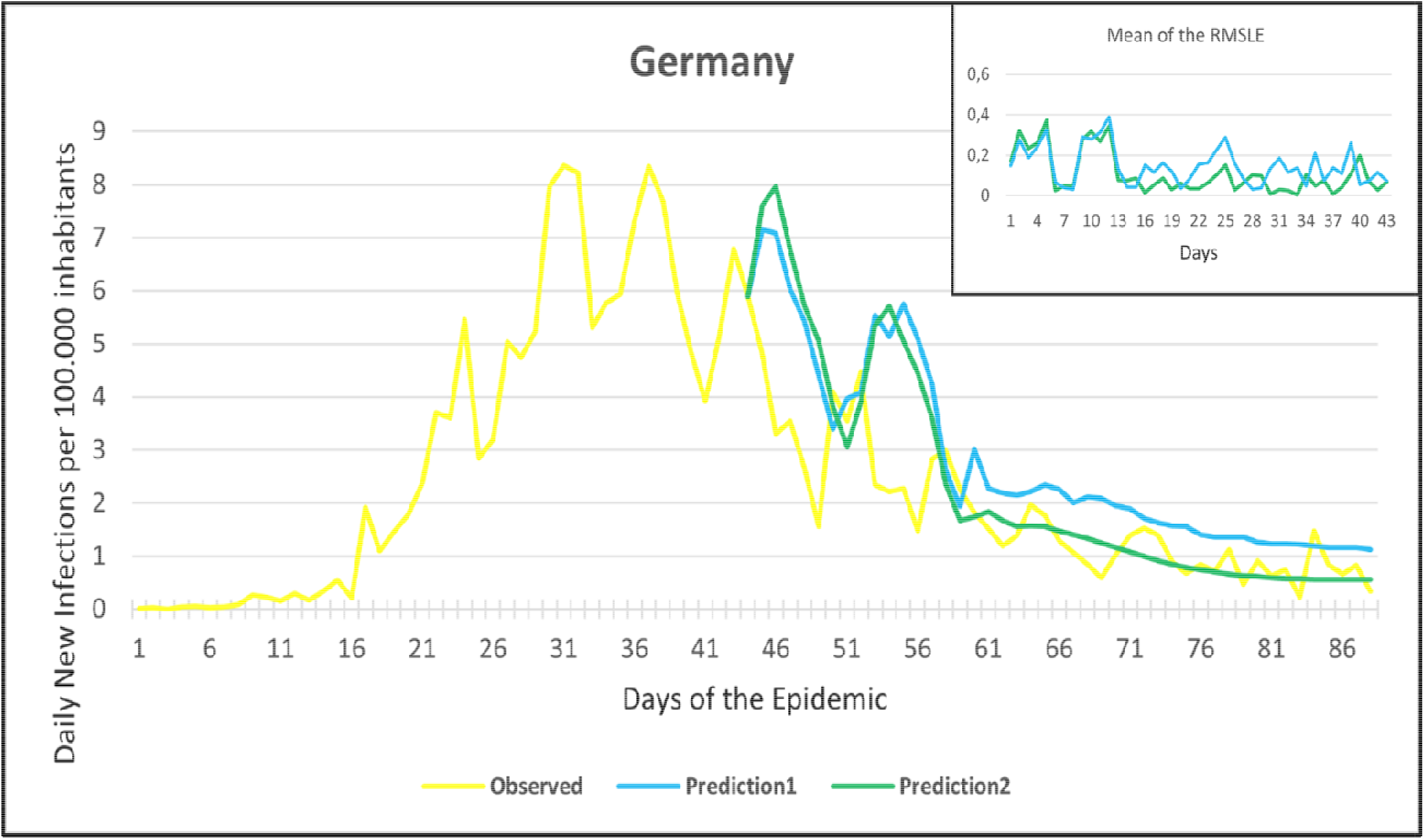
Observation and predictions for the Germany. The small graph in the upper right corner shows the daily error values calculated for the predictions.

**Figure 6.**
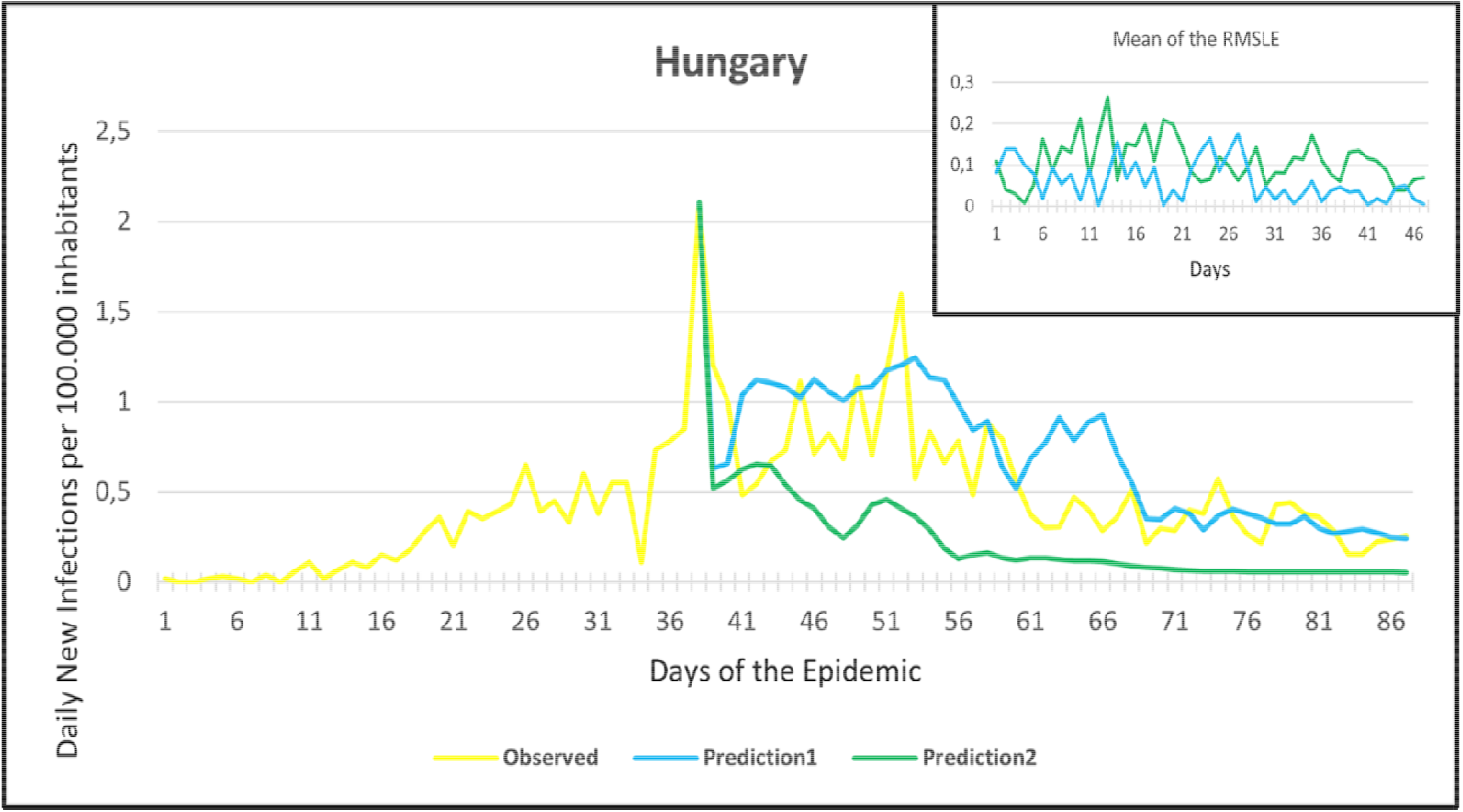
Observation and predictions for Hungary. The small graph in the upper right corner shows the daily error values calculated for the predictions.

**Figure 7.**
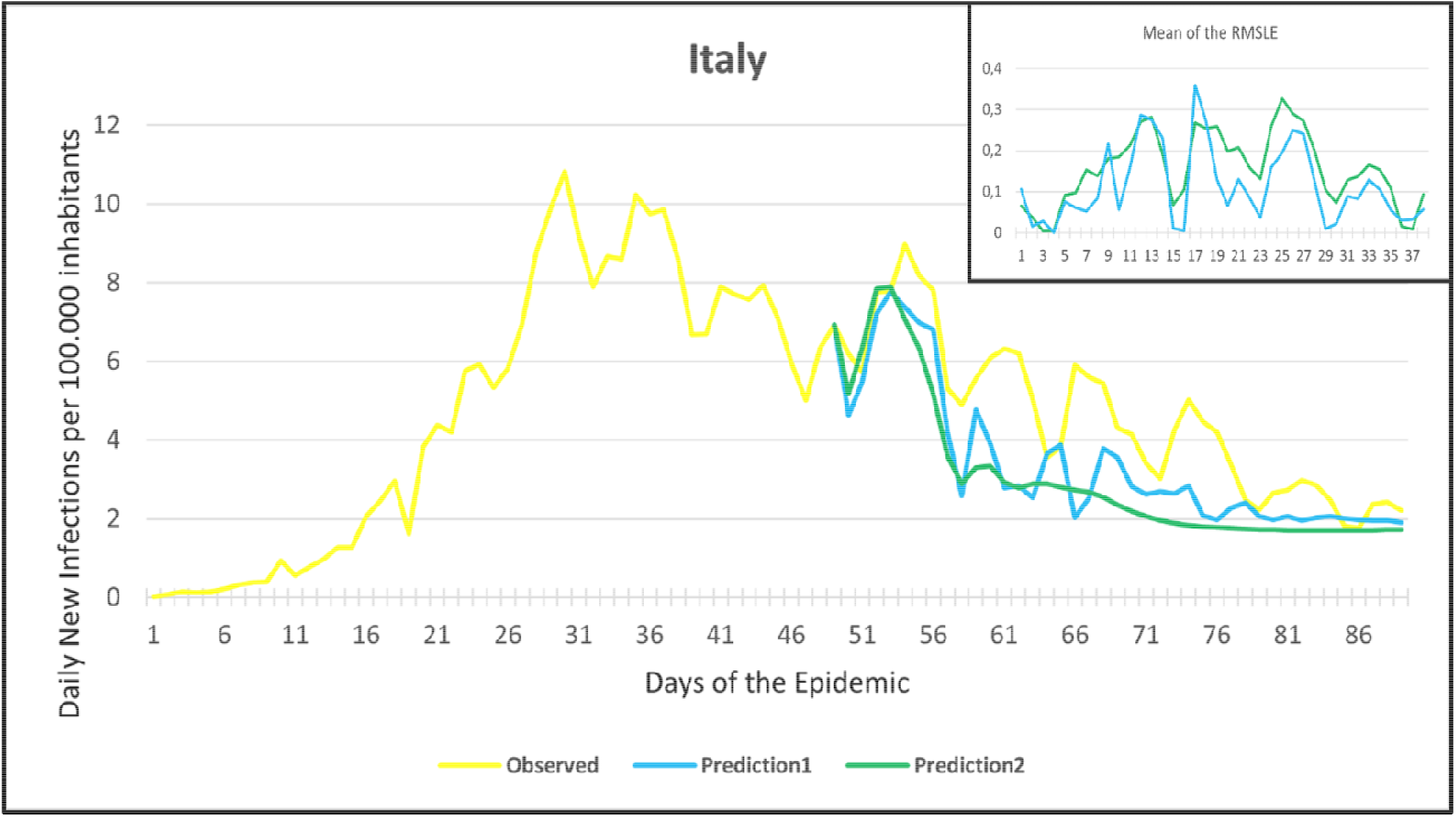
Observation and predictions for Italy. The small graph in the upper right corner shows the daily error values calculated for the predictions.

**Figure 8.**
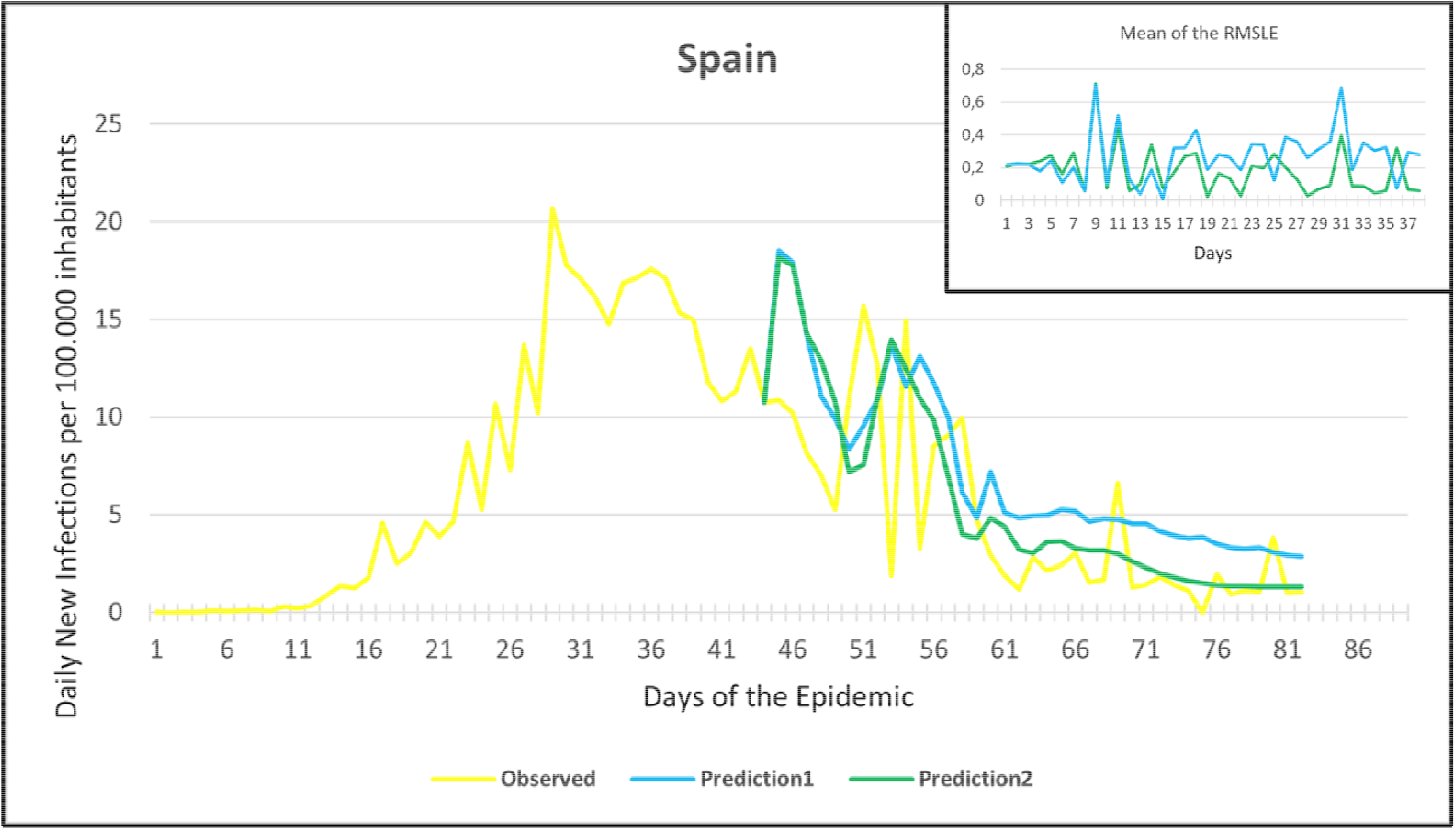
Observation and predictions for Spain. The small graph in the upper right corner shows the daily error values calculated for the predictions.

**Figure 9.**
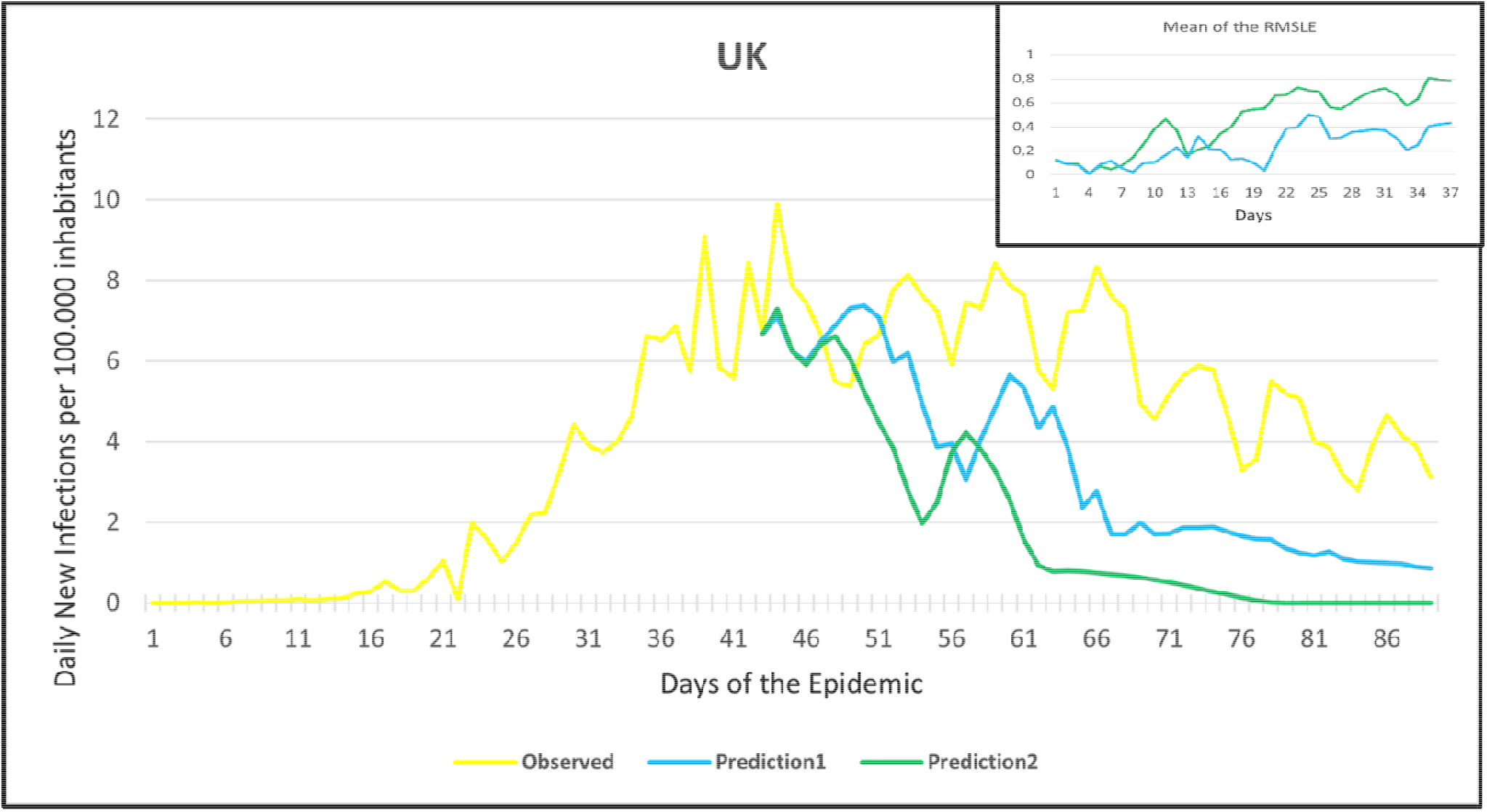
Observation and predictions for the United Kingdom (UK). The small graph in the upper right corner shows the daily error values calculated for the predictions.

**Figure 10.**
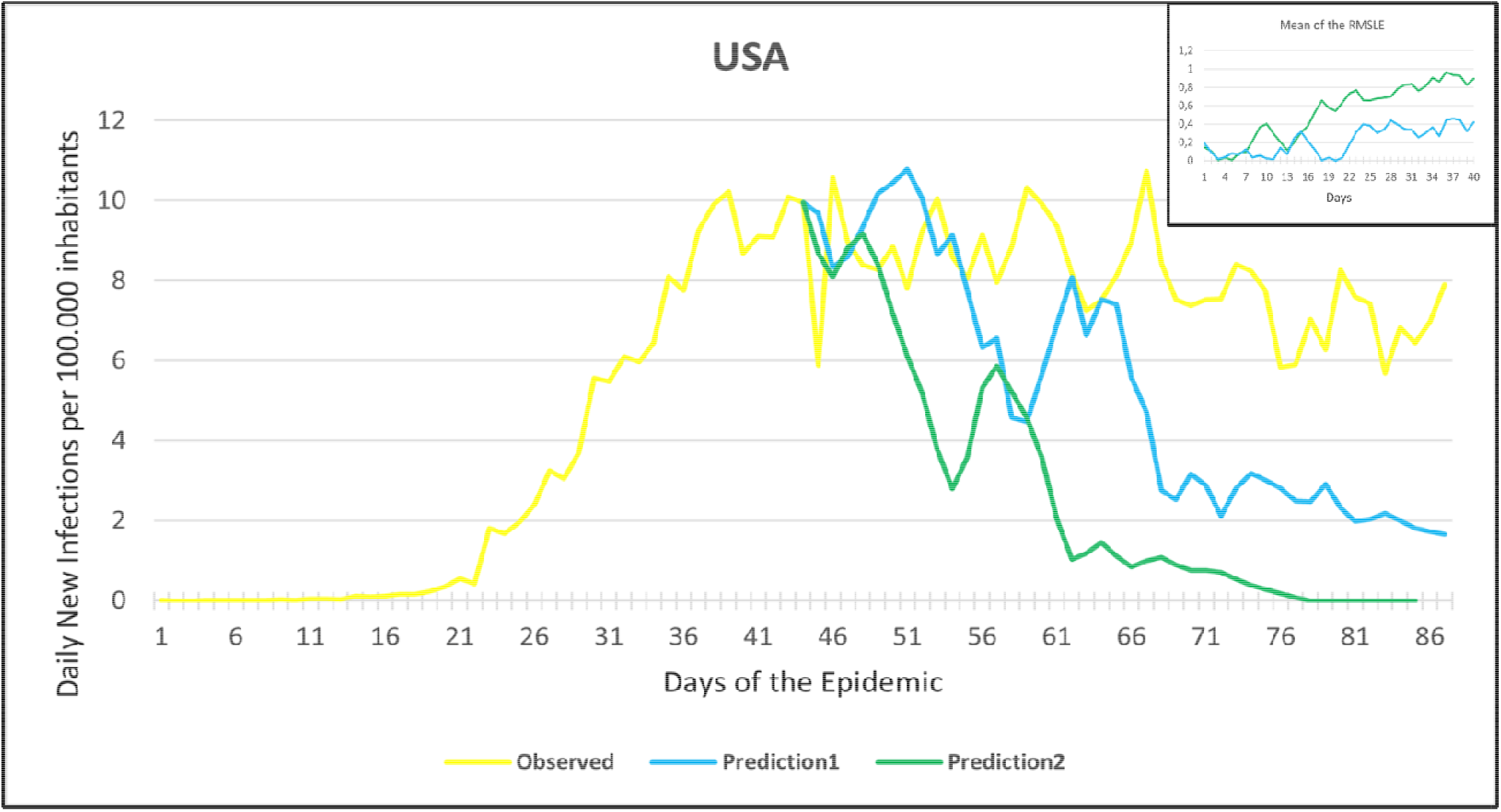
Observation and predictions for the United States of America (USA). The small graph in the upper right corner shows the daily error values calculated for the predictions.

## 4. DISCUSSION

The current COVID-19 pandemic has opened avenues for more effective and efficient application of AI in various aspects of fighting and tackling infectious diseases. At the early stage of the COVID-19 outbreak when little data were available regarding the nature and transmissibility of SARS-CoV-2, modelling studies have attempted to predict the epidemic outcomes using Susceptible-Exposed-Infectious-Recovered (SEIR) model, based on data from Wuhan, China ; the starting point of the outbreak [29]. Besides, forecasting and predicting the COVID-19 trajectories was not the only application of AI during the current pandemic. AI has been adopted in contact-tracing, tracking public health behaviors, and currently in COVID-19 vaccination [30,31]. Since March 2020, we have had the opportunity in our present study to use the declared numbers of daily new cases of COVID-19 to predict our models and to compare the predicted trajectories with the observed data. In countries that imposed strict measures (e.g., strict lockdown), for example, France, Hungary, Italy, and Spain, the predicted models and observed data were closely similar, however, this was not the case of the UK and the USA. This could be attributed to the fact that the learning dataset was based mainly on data from Hubei province where a strict total lockdown was imposed there, unlike the UK and the USA. Besides, we can notice in the models that *Prediction 1* is more accurate for some countries (Hungary, Italy, UK, USA), while *Prediction 2* is more accurate for others (France, Germany, Spain).

The findings of our study underscores that the COVID-19 pandemic is a propagated source outbreak, therefore repeated peaks on the epidemic curve (rise of the daily number of newly diagnosed infections) are to be anticipated. Predictions that were made using AI-based RNNs further implicate that albeit the majority of investigated countries are near or over the peak of the curve, they should prepare for a series of successively high peaks in the near future, until all susceptible people will be infected by the SARS-CoV-2, or effective preventive (e.g., vaccination) or treatment options will become available and utilized effectively. These scenarios are similar to other known propagated source epidemics, e.g., SARS and measles [32].

Albeit suppression and mitigation measures can reduce the incidence of infection, COVID-19 disease, given its relatively high transmissibility reflected by average R0 values of 3.28, will continue to spread, most likely [16]. Accordingly, public health measures must be implemented as the incubation period of the virus may be long (1-14 days, but there are some opinions, that this can be 21 days), during which time asymptomatic or pre-symptomatic spreading may ensue. Moreover, currently it is uncertain, whether those, who were diagnosed with COVID-19 infection, will acquire sufficient immunity or not [13]. Finally, data from countries with warm climates suggest that summer is unlikely to stop the pandemic, as the virus already spreading in Australia and South Africa as well [15,19]. This is why the recurrence of another peak is very likely, and the end of the pandemic cannot be accurately predicted at this time.

Nevertheless, recent publications showed that the earlier the mitigation attempts are in place (e.g., border closure, closing schools, the lockdown of the country, curfew), the more effective is the reduction of the spread of the epidemic [19]. In fact, analyzing the effects of a suppression strategy concerning the COVID-19, it was shown that early implementation of suppression at 0.2 deaths per 100 000 population per week could save 30.7 million lives compared to late implementation of these measures at 1.6 deaths per 100 000 population per week [33]. This seems to be the case in the countries, which had prior knowledge regarding coronavirus infections (e.g., China, Singapore, Hong Kong), as they were more prepared to implement public health measures, had more equipment and health care personnel in place to mitigate the spread of the infections. Those countries, that failed to implement efficient and strict mitigation policies in a timely manner, were facing difficulty in controlling the spread of the disease, as is the case of Italy, the UK and the USA [16].

To the best of our knowledge, this is the first study to model the predicted evolution of the newly diagnosed infections using data from official databases with the help of the AI-based RNNs trained on the currently available data, which were validated by RMSLE calculation. Most studies to date expect a single peak of the epidemic curve, but some fear the emergence of future peaks when mitigation-suppression measures will be discontinued. According to our model, this can even happen, if strict measures are sustained.

Nevertheless, the are some limitations in our study. As the nature of SARS-COV-2 is relatively unknown or dynamic, and it is prone to mutations, the prediction of the spread of the pandemic is not an easy mission. Factors that influenced the reported new cases per day, for example, the efficiency of reporting, the different quality and timing of public health measures, country-specific age-pyramid, and chronic disease burden of the population were not included in our training data set due to lack of reliable data. We did not investigate the number of deaths and recoveries, as we found no reliable data at that time. Similarly, the data regarding diagnostic tests performed per country, or death rates were omitted, given they are highly influenced by the countries’ economic wellbeing, health care systems, facilities and capacities, and other factors [34,35]. There are lots of unforeseen uncertainties and coincidences which could not be implemented in our model, for example, there were days when a large number of people have been diagnosed with COVID-19 on one day (for example in care homes in France or Hungary) that caused a large increase in the number of the daily new cases [16].

To summarize, the COVID-19 disease is a global health challenge, which forced the WHO to declare it a “public health emergency of international concern on 30/01/2020” and later as a global pandemic [18]. The influence of this global epidemic has dug deep into the day-to-day conduct of everyone, with unforeseen challenges still pending for governments and policymakers. Starting from this, everyone, especially decision-makers must be aware, that the current situation might be just the beginning, and even if strict public health measures are executed and sustained, future peaks of infections are possible.

## 5. CONCLUSION

The findings of our study underscore that the COVID-19 pandemic is a propagated source epidemic, therefore repeated peaks of the rise of the daily number of newly diagnosed infections are to be anticipated. In countries where strict control measures were imposed, the predicted models were closely similar to the observed data. Most studies to date expect a single peak on the epidemic curve, but some fear the emergence of future peaks when mitigation-suppression measures will be discontinued. According to our models, this can even happen, if strict measures are sustained. The AI-based predictions might be useful tools and can be recalculated according to the newly observed data to get a more precise forecast of the pandemic. Finally, AI-based predictions are expected to provide public health practitioners and decision makers with sufficient data that would be useful in improving countries’ preparedness to the next stage of a pandemic.

## Data Availability

All data sources are publicly available and described in the methods section.

## Author Contributions

Conceptualization, LRK, TB, JZ, AH; methodology, the methodology was discussed by all co-authors; Data curation: AH, ABA, TB, AT, IV led the data curation and formal analysis; TB, ABA, and GJS made the data extraction and updating. The AI-based analysis and validation was made by AT, TB, IV, AH. Visualizations were made by AT, TB, IV, GJS, AH, ABA, and LRK. Writing—original draft preparation, all authors; Writing—review and editing, all authors; funding acquisition, JZ, and AH. The literature search was done by LRK, ABA, AH, SH, GJS, SG, JZ, RG. The interpretation and review of the literature, methods, results were made in close collaboration by all co-authors. All authors have read and agreed to the submitted version of the manuscript.

## Institutional Review Board Statement

Not Applicable.

## Informed Consent Statement

Not Applicable

## Data Availability Statement

All data sources are publicly available and were described and cited in the methods section.

## Acknowledgments

ABA is supported by the Tempus Public Foundation’s Scholarship at the University of Debrecen. Also, the authors would like to provide their thanks and appreciation to the World Health Organization and Johns Hopkins University for the publicly available data.

